# Antibodies to *Aedes aegypti* D7L salivary proteins as a new serological tool to estimate human exposure to *Aedes* mosquitoes

**DOI:** 10.1101/2023.12.22.23300438

**Authors:** Sophana Chea, Laura Willen, Sreynik Nhek, Piseth Ly, Kristina Tang, James Oristian, Roberto Salas-Carrillo, Aiyana Ponce, Paola Carolina Valenzuela Leon, Dara Kong, Sokna Ly, Ratanak Sath, Chanthap Lon, Rithea Leang, Rekol Huy, Christina Yek, Jesus G. Valenzuela, Eric Calvo, Jessica E. Manning, Fabiano Oliveira

## Abstract

1

**Introduction:** *Aedes spp.* are the most prolific mosquito vectors in the world. Found on every continent, they can effectively transmit various arboviruses, including the dengue virus which continues to cause outbreaks worldwide and is spreading into previously non-endemic areas. The lack of widely available dengue vaccines accentuates the importance of targeted vector control strategies to reduce the dengue burden. High-throughput sensitive tools to estimate human-mosquito contact and evaluate vector control interventions are lacking. We propose a novel serological tool that allows rapid screening of large human cohorts for exposure to potentially infectious mosquitoes and effective targeting of vector control.

**Methods:** We tested 563 serum samples from a longitudinal pediatric cohort study previously conducted in Cambodia. Children enrolled in the study were dengue-naïve at baseline and were followed biannually for dengue incidence for two years. We used Western blotting and enzyme-linked immunosorbent assays to identify the most immunogenic *Aedes aegypti* salivary proteins and measure total anti-*Ae. Aegypti* IgG.

**Results:** We found a strong correlation (r_s_=0.86) between the combined IgG responses against AeD7L1 and AeD7L2 recombinant proteins and those to whole salivary gland homogenate. We observed seasonal fluctuations of AeD7L1+2 IgG responses, corresponding to *Aedes spp.* abundance in the region, and no cross-reactivity with *Culex quinquefasciatus* and *Anopheles dirus* mosquitoes. The baseline median AeD7L1+2 IgG responses for young children were higher in those who developed asymptomatic dengue versus those who developed symptomatic dengue.

**Conclusion:** The IgG response against AeD7L1+2 recombinant proteins is a highly sensitive and *Aedes* specific marker of human exposure to *Aedes* bites that can facilitate standardization of future serosurveys and epidemiological studies by its ability to provide a robust estimation of human-mosquito contact in a high-throughput fashion.

## 2 Introduction

Dengue is the most prevalent arboviral disease worldwide, with the greatest burden in tropical and sub-tropical regions (1, 2). Half the world’s population is at risk of contracting dengue, including 1.3 billion people living in ten dengue-endemic countries of the South-East Asian region (3). From 2015 to 2019, dengue cases in the region increased by 46% culminating in a major epidemic in 2019 that led to the highest number of annual global cases (>5.2 million) ever to be reported in the same year (4).

Currently, existing vaccines remain selectively available; thus, prevention and control of dengue heavily rely on effective vector control measures targeted towards *Aedes* mosquitoes. The efficacy of these vector control methods is currently evaluated using expensive and labor-intensive insect trapping that may not reflect true human exposure to mosquito bites. An alternative approach exploits the human humoral immune responses against the vector’s salivary components that are injected during a bloodmeal. Salivary proteins of several hematophagous arthropods were previously identified and shown to facilitate blood feeding by limiting the host’s vasoconstriction, inhibiting coagulation processes, and suppressing pain receptors (5). Saliva of *Aedes aegypti* female mosquitoes contains about 100 abundant salivary proteins (6, 7) that are implicated in enhanced viral dissemination and increased pathogenesis of viral infections (8).

IgG responses against whole salivary gland homogenate of *Aedes* mosquitoes (SGH) correlate with the intensity of mosquito bite exposure (9), and were previously proposed as a marker of human exposure to *Aedes* mosquitoes (10–12). Since then, researchers established a repertoire of promising recombinant salivary antigens to bypass the potential cross-reactivity of SGH with other vector species and to facilitate the use of biomarkers of *Aedes* exposure in large-scale cohort studies (13), with the Nterm-34kDa peptide biomarker being the most studied (13, 14).

Interestingly, none of these studies assessed how well the antibody (Ab) responses against *Ae. aegypti* recombinant proteins correlate with those against whole saliva or SGH, the reference standard for *Aedes* exposure. Ab responses against whole SGH and several recombinant *Ae. aegypti* salivary proteins have also been implicated in associations with disease outcome(15, 16). IgG reactivity to *Ae. aegypti* salivary proteins were higher in DENV positive patients than in uninfected patients (9, 15, 17). Conversely, Ab responses against the *Ae. aegypti* D7 proteins, involved in the scavenging of biogenic amines and cysteinyl leukotrienes and, thus, potentially involved in preventing the host’s inflammatory response, were higher in DENV positive and febrile patients as opposed to non-febrile patients. In the present study, we identified salivary biomarkers of *Ae. aegypti* exposure that can replace the use of mosquito saliva or SGH and subsequently investigate if a relationship exists between these surrogate biomarkers of exposure and clinical outcomes in dengue infected patients in a longitudinal pediatric cohort in Cambodia (18).

## 3 Results

### 3.1 Identification of immunogenic *Aedes aegypti* salivary proteins in a Cambodian pediatric population

We used Western blot to measure the recognition of *Ae. aegypti* salivary proteins by IgG present in the serum of our study population. We observed sera reactivity in the *Ae. aegypti* SGH molecular weight range of 28 kDa to 98 kDa (Fig 1A). Cambodian sera reactivity increased on par with anti-SGH ELISA OD values (Fig. 1A, Supplementary Fig. 1). Using a digital detection system, we identified reactive protein bands with approximate molecular weights of 80 kDa, 62 kDa, 35 kDa and 30 kDa and accounting for 72 %, 88 %, 85 %, and 82 % of the total seropositivity, respectively. We next produced and screened 18 *Ae. aegypti* recombinant salivary proteins by Western blot using a pool of *Ae. aegypti* SGH reactive sera from our cohort (n=10) (Fig. 1C). We identified five of the 18 *Ae. aegypti* recombinant salivary proteins as reactive, corresponding to AeApyrase, AeD7L1, AeD7L2, NIH-23, and NIH-27 (Table 1, Supplementary Fig. 2). We scaled up production of these five *Ae. aegypti* salivary recombinant proteins, verified their purity by Coomassie G-250 staining (Fig. 1B) and tested their reactivity to the pooled sera (Fig. 1C). Importantly, sera from healthy U.S. blood donors who were non-reactive on an *Ae. aegypti* SGH ELISA, also did not react to any of the tested recombinant proteins (Fig. 1C).

**Figure 1.**
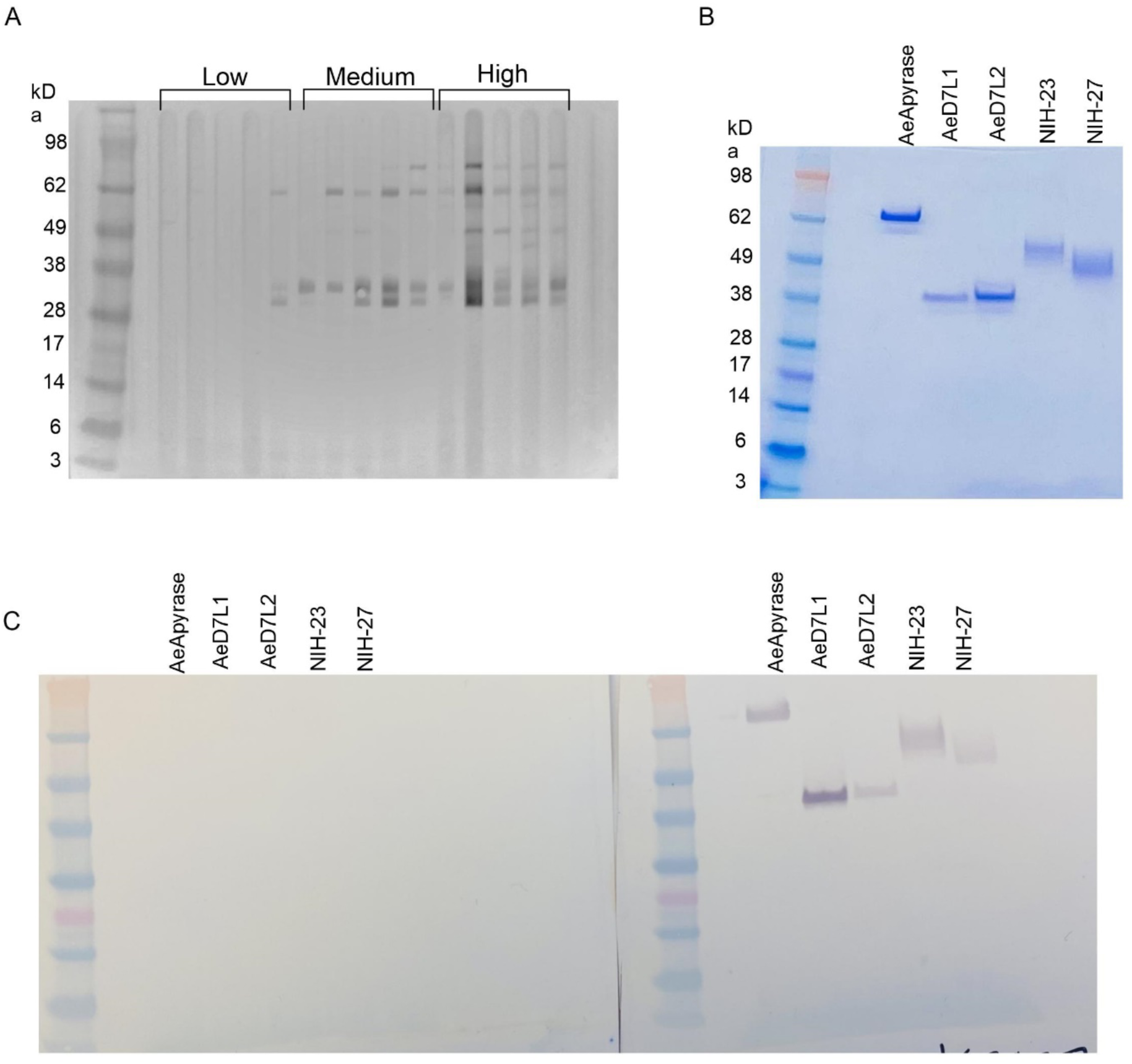
Sera from children living in dengue-endemic areas in Kampong Speu province, Cambodia recognize native SGH from *Ae. aegypti*. (A) Western blot displaying reactivity of Cambodian sera samples organized by increasing *Ae. aegypti* SGH ELISA OD values. (B) Coomassie blue stained SDS-PAGE of the five selected recombinant proteins (AeD7L2, AeD7L1, NIH-27, NIH-23, and AeApyrase). (C) Western blot with AeD7L2, AeD7L1, NIH-27, NIH-23, and AeApyrase against a pool of negative (left) and positive (right) sera.

**Table 1.**
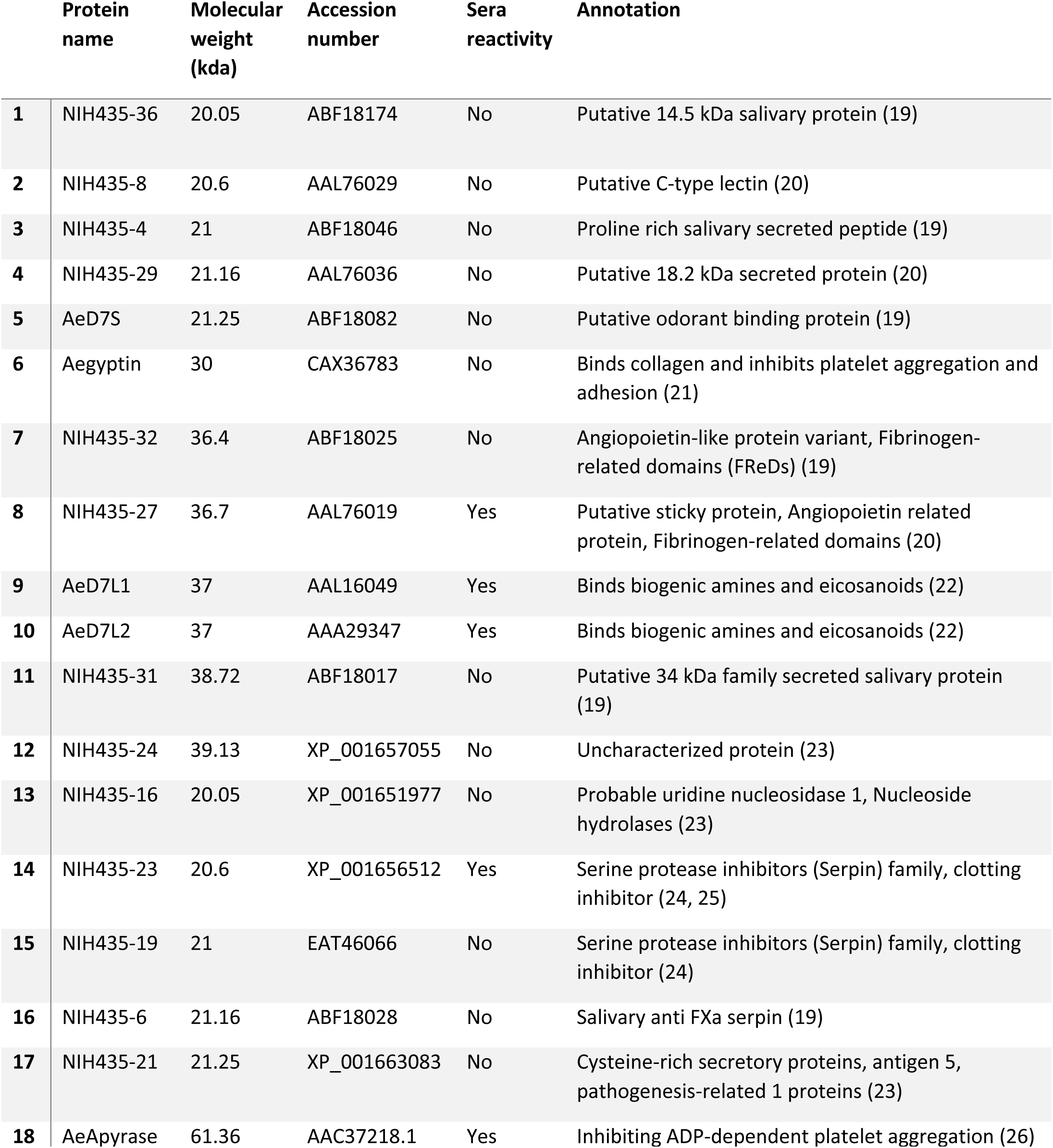
Name, molecular weight, accession number, serum reactivity, and function of the 17 recombinant proteins used for the immunoreactivity screening against *Ae. aegypti* salivary gland homogenate.

|  | Protein name | Molecular weight (kda) | Accession number | Sera reactivity | Annotation |
| --- | --- | --- | --- | --- | --- |
| 1 | NIH435-36 | 20.05 | ABF18174 | No | Putative 14.5 kDa salivary protein (19) |
| 2 | NIH435-8 | 20.6 | AAL76029 | No | Putative C-type lectin (20) |
| 3 | NIH435-4 | 21 | ABF18046 | No | Proline rich salivary secreted peptide (19) |
| 4 | NIH435-29 | 21.16 | AAL76036 | No | Putative 18.2 kDa secreted protein (20) |
| 5 | AeD7S | 21.25 | ABF18082 | No | Putative odorant binding protein (19) |
| 6 | Aegyptin | 30 | CAX36783 | No | Binds collagen and inhibits platelet aggregation and adhesion (21) |
| 7 | NIH435-32 | 36.4 | ABF18025 | No | Angiopoietin-like protein variant, Fibrinogen-related domains (FReDs) (19) |
| 8 | NIH435-27 | 36.7 | AAL76019 | Yes | Putative sticky protein, Angiopoietin related protein, Fibrinogen-related domains (20) |
| 9 | AeD7L1 | 37 | AAL16049 | Yes | Binds biogenic amines and eicosanoids (22) |
| 10 | AeD7L2 | 37 | AAA29347 | Yes | Binds biogenic amines and eicosanoids (22) |
| 11 | NIH435-31 | 38.72 | ABF18017 | No | Putative 34 kDa family secreted salivary protein (19) |
| 12 | NIH435-24 | 39.13 | XP_001657055 | No | Uncharacterized protein (23) |
| 13 | NIH435-16 | 20.05 | XP_001651977 | No | Probable uridine nucleosidase 1, Nucleoside hydrolases (23) |
| 14 | NIH435-23 | 20.6 | XP_001656512 | Yes | Serine protease inhibitors (Serpin) family, clotting inhibitor (24, 25) |
| 15 | NIH435-19 | 21 | EAT46066 | No | Serine protease inhibitors (Serpin) family, clotting inhibitor (24) |
| 16 | NIH435-6 | 21.16 | ABF18028 | No | Salivary anti FXa serpin (19) |
| 17 | NIH435-21 | 21.25 | XP_001663083 | No | Cysteine-rich secretory proteins, antigen 5, pathogenesis-related 1 proteins (23) |
| 18 | AeApyrase | 61.36 | AAC37218.1 | Yes | Inhibiting ADP-dependent platelet aggregation (26) |

### 3.2 The combined antibody responses against *Aedes aegypti* D7L1 and D7L2 salivary proteins is a valid surrogate for antibodies against *Ae. aegypti* SGH

We tested a random subset of 18 Cambodian children sampled at seven consecutive visits during the dry and wet seasons from 2018 to 2021 on an ELISA assay for Abs against the selected recombinant proteins or against SGH (Fig. 2A). We achieved a high correlation between SGH and AeD7L1+2 (r = 0.91, r_s_ = 0.92), AeD7L1 (r = 0.82, r_s_ = 0.82), and AeD7L2 (r = 0.76, r_s_ = 0.82), alone or when combined with AeApyrase, AeNIH-27 or AeNIH-23 (Fig. 2). No significant correlation was observed between Abs against SGH and a negative control antigen (Bovine Serum Albumin, BSA) (r = 0.17 (*P* = 0.05), r_s_ = 0.15 (*P* = 0.1)). We selected the combination of AeD7L1+2 as the optimal and most parsimonious candidate for further analyses. All *P* values and confidence intervals are displayed in Supplementary Table 1.

**Figure 2.**
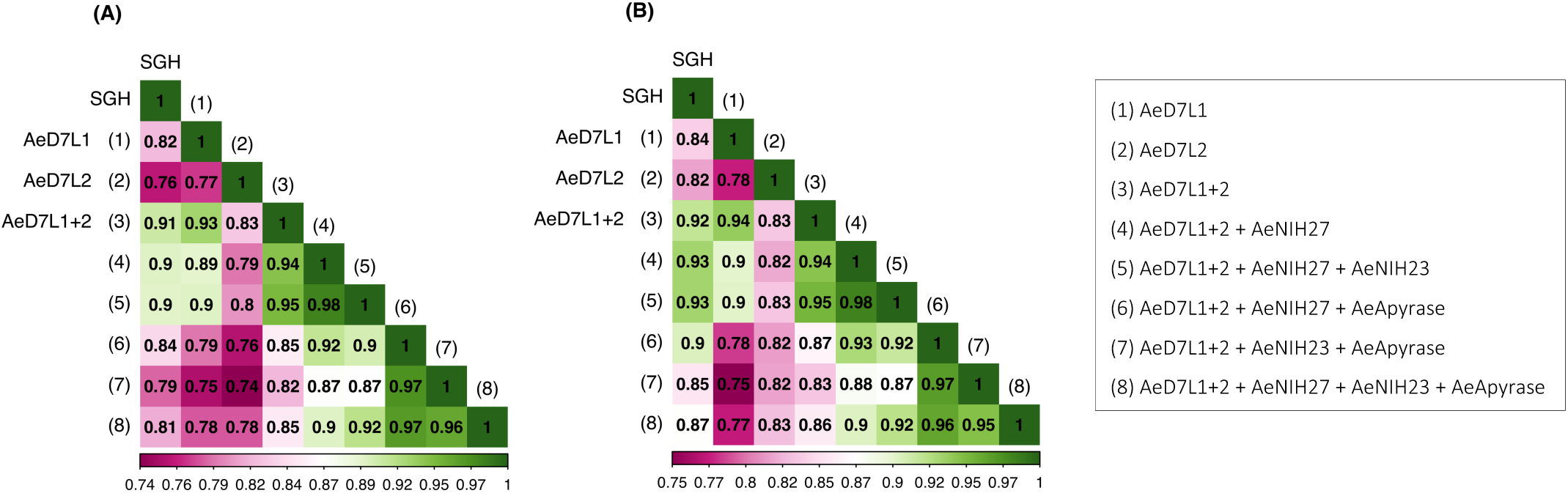
Heat map showing the Pearson **(A)** and Spearman **(B)** correlation coefficients (r and rs, respectively) between antibody responses against SGH and against AeD7L1+2, AeD7L1, or AeD7L2, alone or when combined with AeApyrase, AeNIH-27 or AeNIH-23.

### 3.3 Antibody responses show a seasonal fluctuation that is most pronounced for AeD7L1 and AeD7L1+2

Wet and dry seasons in Cambodia reflect the seasonal abundance of *Aedes spp.* We detected significant differences in anti-SGH, anti-AeD7L1, anti-AeD7L2, and anti-AeD7L1+2 IgG levels between wet and dry seasons for the same subset of 18 Cambodian children who were sampled at seven consecutive visits (Fig. 3). More so, the seasonal fluctuation for AeD7L1 and the composite marker AeD7L1+L2 was stronger (moderate effect size, Kendall W = 0.33 and 0.30, respectively) than that for SGH or AeD7L2 (small effect size, Kendall W=0.17 and 0.19, respectively). No significant differences were observed for the Ab responses against BSA during the wet and dry seasons (Supplementary Fig. 3).

**Figure 3.**
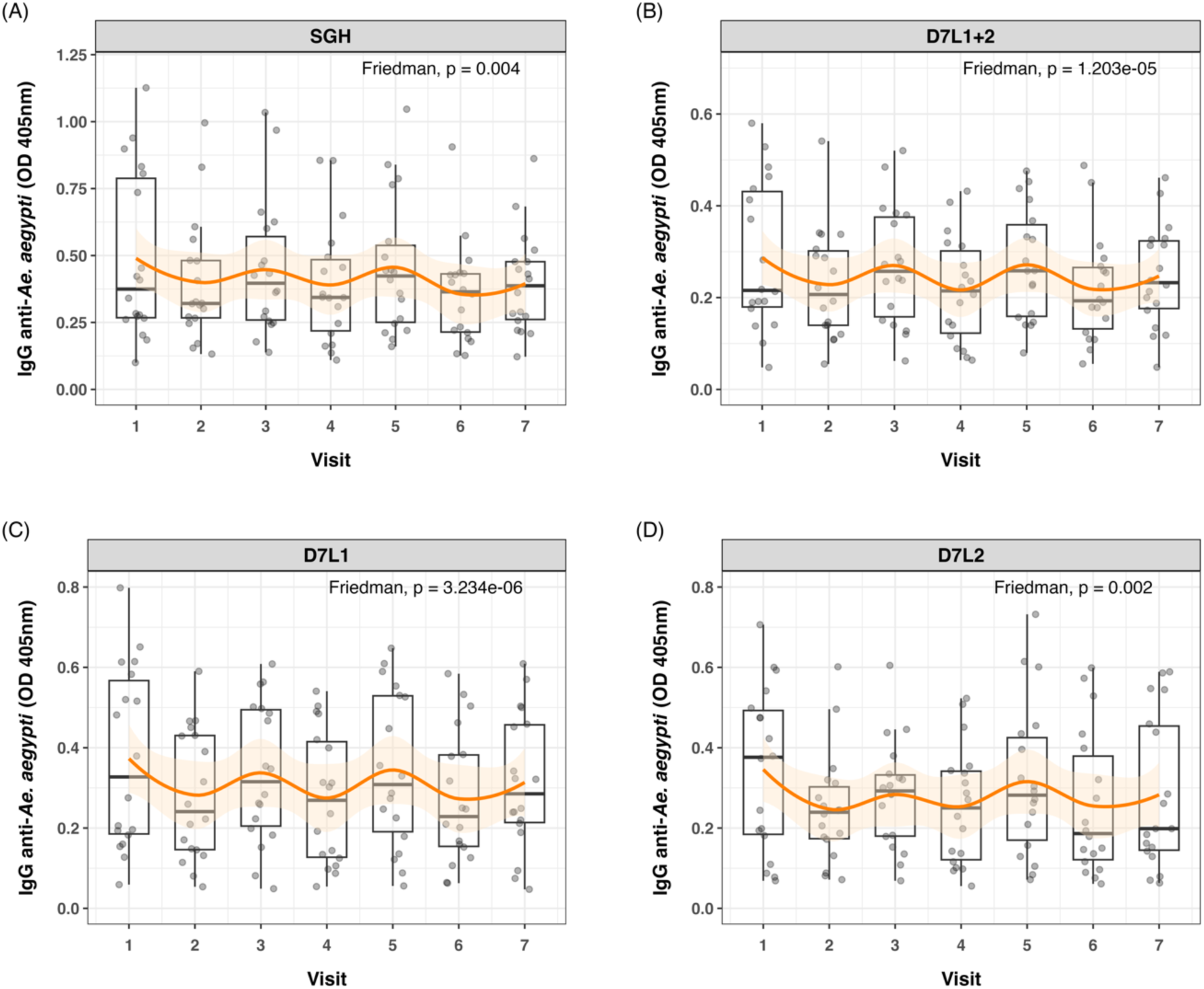
Seasonal variation in anti-*Ae. aegypti* IgG levels. Dot and boxplots of standardized ELISA optical density (OD) values of 18 individuals tested at seven different time points (Visit 1 to 7) for IgG against (A) SGH, (B) AeD7L1+2, (C) AeD7L1, and (D) AeD7L2. Visit 1, 3, 5, and 7 took place in wet seasons from 2018 to 2021, the other visits took place during the dry seasons. The solid black horizontal line within the boxplots is the median; the lower and upper borders are, respectively, the 1st and 3rd quartiles; the vertical bars indicate the minimum and maximum values. The orange smoothed fitted curves represent a loess regression (span α = 0.45) with 95% confidence intervals (shaded area). Results of the overall Friedman rank test are provided.

### 3.4 Children who developed asymptomatic dengue had higher anti-AeD7L1+2 antibody titers at baseline than those who developed symptomatic dengue

Sera samples of all children naïve at baseline (n=563) were tested for the response against AeD7L1+2 to assess associations with disease outcome (i.e., no dengue infection, asymptomatic dengue, and symptomatic dengue). Correlation coefficients between *Ae. aegypti* SGH and the AeD7L1+2 recombinant proteins remained strong for the full cohort (r=0.85, r_s_=0.86). To investigate associations with anti-*Aedes* Ab levels at baseline and disease, we first fitted Cox PH models that consider time to any dengue infection. We did not observe any significant differential risk for dengue between children with high or low baseline Ab levels, for neither SGH, nor AeD7L1+2 (Supplementary Table 2 and 3). We then tested the distribution of baseline Ab responses to SGH and AeD7L1+2 with age and disease outcome group. Accounting for age, disease outcome, and an age x disease outcome interaction term, multivariate models revealed that overall Ab responses decreased with increasing age for both SGH and D7L1+2 (Fig. 4A). However, the Ab response of children who developed symptomatic dengue increased with age compared to children who developed asymptomatic dengue (test of age × disease outcome interaction term (Fig. 4A). Marginal effect plots of the age x disease outcome interaction from the GLM regression indicate a cross-over in baseline Ab responses at six years old (Fig. 4B). Indeed, when comparing median baseline Ab levels between disease outcomes, a significant difference existed among children younger than six years of age who developed asymptomatic vs. symptomatic dengue, for both SGH and AeD7L1+2 (P = 0.046 and P = 0.022, respectively, Fig. 4C). Children older than six years did not exhibit differences in baseline median Ab responses between disease outcome groups (Fig. 4C).

**Figure 4.**
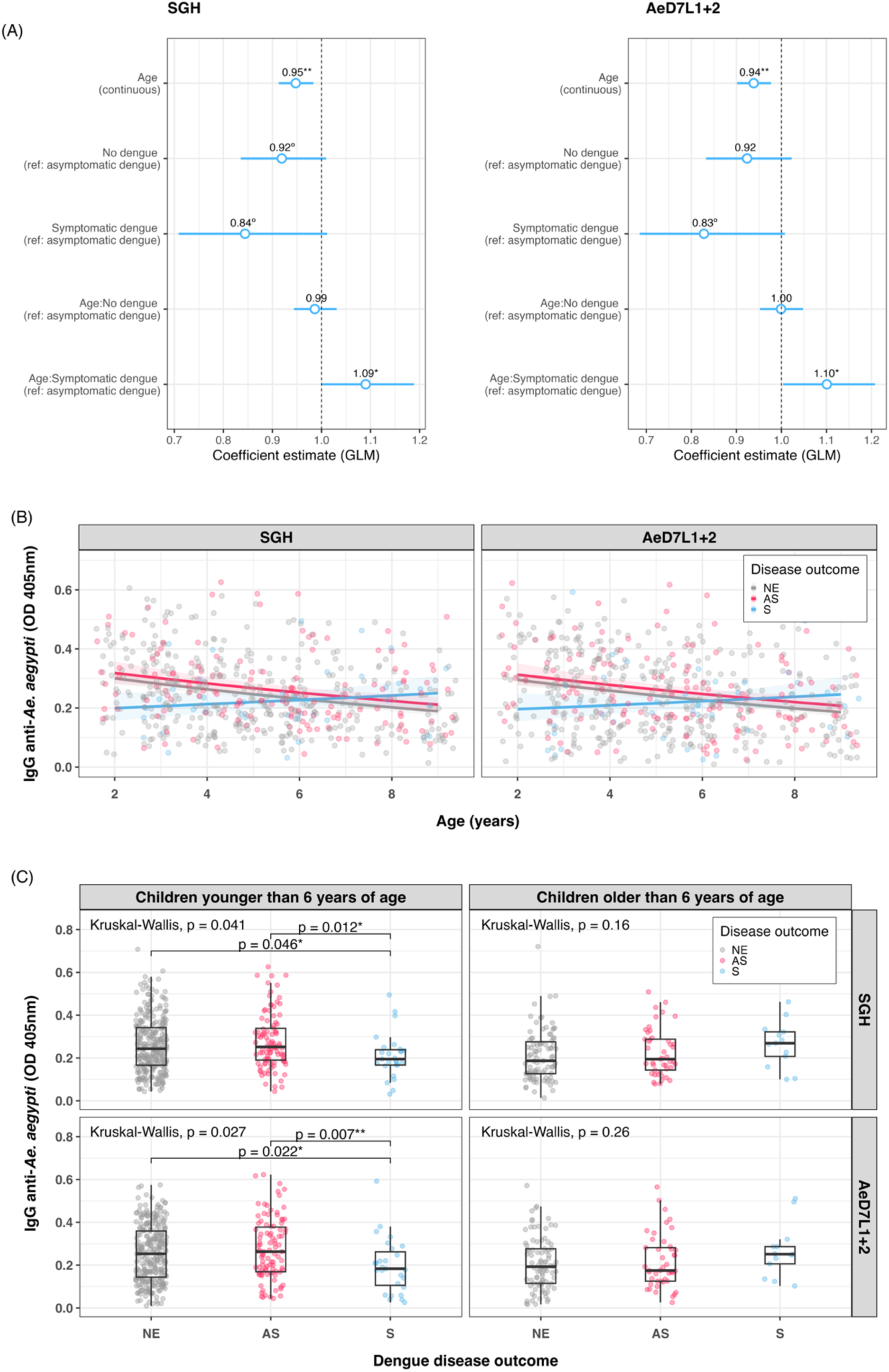
Trends in baseline anti-*Ae. aegypti* IgG responses. (A) Exponentiated regression coefficient estimates for the generalized linear model fitted to the IgG data per assay. The circles represent the exponentiated mean coefficient estimate (the value shown above the corresponding circle). The horizontal lines represent the 95% confidence intervals of the estimate. The referent group was “asymptomatic” for both disease outcome and the age × disease outcome interaction term. (B) Scatterplot of standardized ELISA IgG optical density (OD) values with age at study baseline, for both SGH and AeD7L1+2 (n=563). The solid lines visualize the marginal effect of the age x disease outcome interaction from the GLM regression fitted to the antibody data. (C) Dot and boxplots of IgG OD values for SGH and AeD7L1+2, comparing different dengue disease outcomes for children younger or older than six years old. The solid black horizontal line within the boxplots is the median; the lower and upper borders are, respectively, the 1st and 3rd quartiles; the vertical bars indicate the minimum and maximum values. Results of the overall Kruskal-Wallis test and the post-hoc Dunn’s test for pairwise comparisons are provided. GLM = Generalized Linear Model; OD = Optical Density; NE = No Event; AS = Asymptomatic dengue; S = Symptomatic dengue; SGH = Salivary Gland Homogenate. Significance levels *** *P*<0.001; ** *P*<0.01; * *P*<0.05; ° *P*<0.1.

### 3.5 Antibody reactivity to AeD7L1+2 recombinant salivary proteins is specific to *Aedes* mosquitoes

Since Cambodians are exposed to the bites of several hematophagous insects during their life, we tested for potential cross-reactions between AeD7L1+2 proteins and salivary proteins from other mosquito species abundant in our study area using an inhibition immunoblot assay. We incubated SGH from *Ae. albopictus*, *Cx. quinquefasciatus*, and *An. dirus* with a pool of Cambodian sera reactive to *Ae. aegypti* SGH and observed recognition of several salivary proteins at diverse molecular weights, including the ones respective to AeD7L1+2 (Fig 5A, left panel). We then incubated the sera pool with AeD7L1+2 recombinant proteins, to absorb AeD7L1+2 specific Abs. The sera reactivity at 28-38 kDa (corresponding to the MW of AeD7L1 and AeD7L2 proteins) was lost for both *Ae. aegypti* and *Ae. albopictus* SGH (Fig 5A, right panel). However, the sera remained reactive against a *Cx. quinquefasciatus* SGH protein of equivalent molecular weight, suggesting that Cambodian children have anti-*Culex* Abs that do not cross-react with AeD7L1+2. This further supports the notion that AeD7L1+2 recombinant proteins are specific to *Aedes* mosquitoes. Indeed, multiple pairwise alignment of the D7L proteins from *Ae. aegypti*, *Ae. albopictus* and *Cx. quinquefasciatus* shows a high degree of divergence between species with few conserved areas of identity (Fig. 5B, dark shaded areas). At the *Aedes* genus, the AeD7L2 protein sequence is 67.85 % identical to the AlboD7L1 sequence and the AeD7L1 sequence is 71.62 % identical to the AlboD7L2 sequence. Interestingly, AeD7L1 and AeD7L2 protein sequences are only 38.71 % identical. Similarly, the phylogenetic analysis of the Culicidae confirms clustering of AeD7L1 with AlboD7L2, whereas AeD7L2 sequence clustered with AlboD7L1. Proteins from both *Aedes* species, however, were segregated from *Culex* D7L or *Anopheles* D7L proteins.

**Figure 5.**
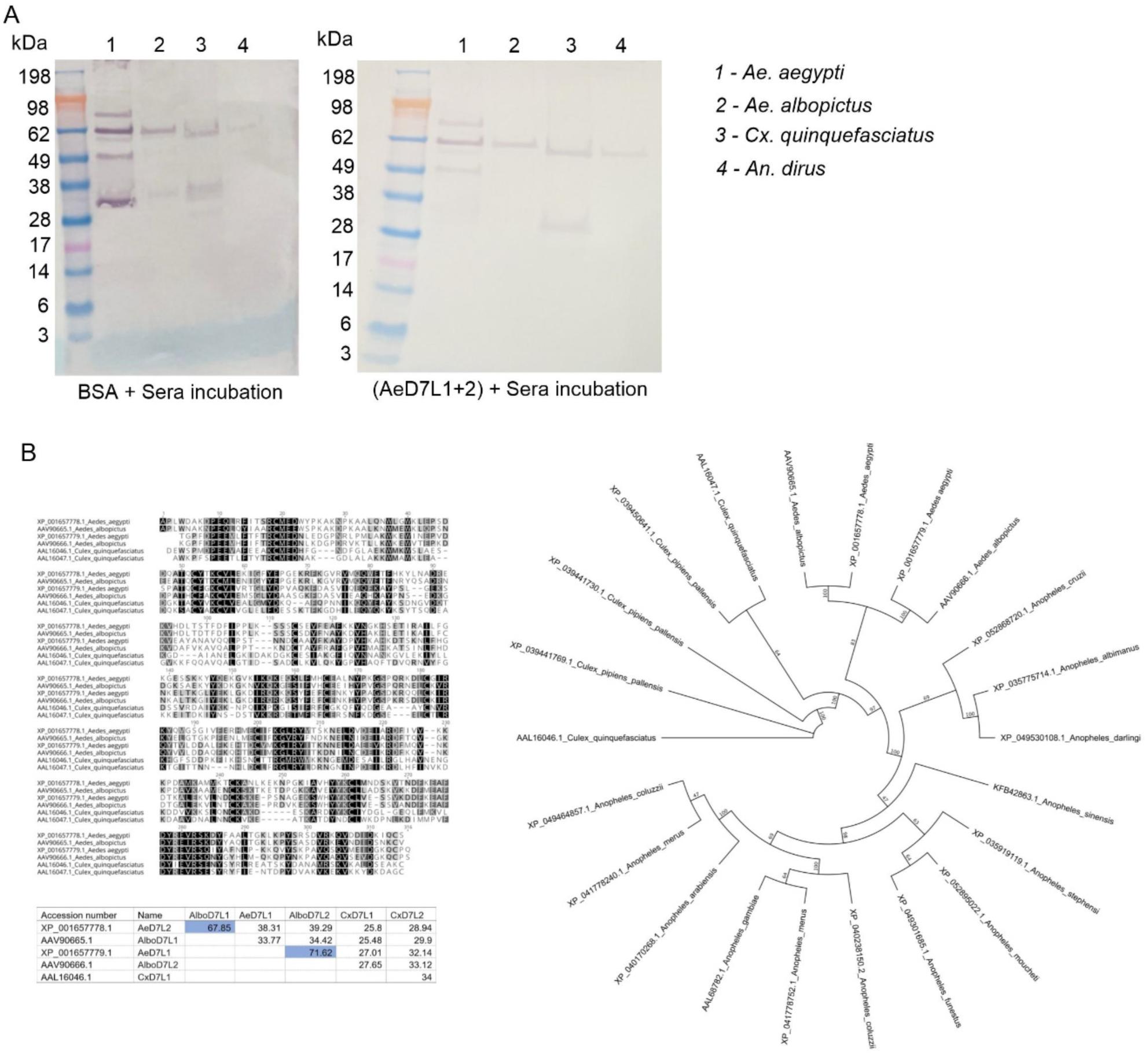
Specificity of AeD7L1+2 recombinant proteins to *Aedes* mosquitoes. (A) Western blots show reactivity of Cambodian sera to SGH of *Ae. aegypti*, *Ae. albopictus*, *Cx. quinquefasciatus*, and *An. dirus* without (left) and with (right) pre-incubation with AeD7L1+2 recombinant proteins. (B) Multiple alignments of D7L proteins from *Ae. aegypti*, *Ae. albopictus*, and *Cx. quinquefasciatus* using MUSCLE. The matrix indicates the percentages of identities between D7L proteins of different species. Color shading indicate percentage identity between sequences, going from 100 % (black) to 0 % identity (white). The evolutionary history was inferred using the Maximum Likelihood method. Evolutionary analyses were conducted in Geneious using the Paup plugin.

## 4 Discussion

*Ae. aegypti* SGH ELISAs are a useful tool to measure mosquito exposure (27), but are costly, laborious, and challenging to standardize between laboratories. Here, we propose an ELISA to measure Ab responses against recombinant *Ae. aegypti* D7L1+2 salivary proteins as a surrogate for monitoring human exposure to *Ae. aegypti* bites. Our hypothesis relies on measuring a rapid reduction of specific IgG Ab levels to AeD7L1+2 when exposure to *Ae. aegypti* is disrupted. *Aedes aegypti* abundance and dengue incidence are known to fluctuate throughout seasons, as has been evidenced by entomological indices and previous serosurveys (28, 29). As a proof of principle, the AeD7L1+2 assay uncovered seasonal weather patterns by capturing the waxing and waning of IgG Abs levels from dry to rainy season. This assay This assay utilizes recombinant proteins, thereby forgoing reliance on insectary maintenance and manual salivary gland dissection and enabling standardization through a can be performed in a high throughput and cost-effective platform and it could be assessed through a simple finger prick. It can be employed to identify hot spots areas with high biting rates (30) or evaluate the effectiveness of vector control methods (31–33), which could then inform allocation of scarce public health resources to areas with a high risk of *Aedes*-borne disease transmission. More so, having an accurate estimate of population exposure to *Aedes* mosquitoes will help raise community awareness and might encourage residents in high-risk areas to take protective measures (27).

The D7 subfamily of salivary proteins are widely present across the order of bloodsucking Diptera (34) and likely regulate hemostasis by scavenging biogenic amines and leukotrienes, which are small molecules that play essential roles in inflammation, allergy, vascular permeability, and vascular tone (35). Indeed, salivary AeD7L knock-out mosquitoes required longer probing times than WT *Ae. aegypti*, confirming its importance on blood feeding (36). The D7L is known to be immunogenic in humans (37) and may serve as potential exposure markers for *An. gambiae* (38). Entomological parameters estimating *Ae. aegypti* density are associated with SGH Ab responses substantiating its use as a reference standard for the identification of new exposure markers (9). For the first time, this study formally correlates side-by-side paired responses to distinct *Ae. aegypti* salivary proteins to the whole *Ae. aegypti* SGH. Our unbiased screening approach provides the confidence that AeD7L1+2 assay is well suited to substitute *Ae. aegypti* SGH in ELISAs. The Abs to AeD7L1+2 seem to recognize both *Ae. aegypti* and *Ae. albopictus*, serving as an *Aedes* generic exposure tool with no detected cross-reactivity to *Cx. quinquefasciatus* or *An. dirus* mosquitoes SGH, a likely advantage to SGH assays, where the risk of cross-reactivity may be higher. These reactivity patterns are explained by the high sequence identity of D7L salivary proteins within the *Aedes* genus and their high degree of divergence compared to D7L salivary proteins from *Culex* or *Anopheles* species. Deploying this *Aedes* sp. tool can be advantageous for arboviral studies since both *Ae. albopictus* and *Ae. aegypti* effectively transmit many of the same viruses (39, 40). In certain occasions, however, specific information may be required to evaluate if the focus of transmission is more urban (*Ae. aegypti*) or rather in forested areas (*Ae. albopictus)*. Our AeD7L1+2 assay could then be complemented with novel species-specific, yet less sensitive, specific salivary protein ELISAs.

Using our three-year longitudinal pediatric cohort study (41), we show that IgG Ab levels against AeD7L1+2 are higher in naïve children younger than six years who developed asymptomatic (primary) dengue than those who developed symptomatic (primary) dengue. This observation may allude to a protective effect of AeD7L Abs in dengue disease outcomes. However, we were unable to replicate this association when including person-time at-risk in the analysis, highlighting the complexity of this “DENV – *Aedes* salivary proteins – anti-saliva Abs” triad. It has been shown that DENV-infected individuals have significantly higher Ab titers against total *Ae. aegypti* SGH than controls [3; 7; 12; 35]. Similarly, IgG levels against the Nterm-34 kDa and AgBR1 were higher in dengue patients without warning signs than in those with dengue with warning signs [10]. However, Abs against the Nterm-34kDa peptide were not able to retrospectively identify individuals who developed DENV infection (14) and in Colombia no statistically significant differences were found when comparing antibodies to D7L between febrile DENV-negative, non-febrile DENV-negative or DENV-positive subjects (17), underscoring the uniqueness of each salivary protein and their distinct role in disease transmission. In the mosquito salivary glands, AeD7L abundance is upregulated during DENV infection and AeD7L is believed to bind DENV directly forming proteins complexes (42). Interestingly, AeD7L inhibited DENV infection in a human monocyte cell line and in vivo in a mouse model (42). The role of AeD7L1+2 Abs in infection adds further complexity to the DENV establishment in humans from endemic areas. AeD7L1+2 Abs may bind to D7 during transmission and help neutralize D7-DENV complexes leading to reduction in viral dissemination and primary asymptomatic dengue. However, the neutralization capacity and the quality of AeD7L1+2 Abs from our dengue asymptomatic versus symptomatic subjects on D7-DENV complexes remains to be tested.

We acknowledge that more data are needed to validate the performance of the AeD7L1+2 assay in other populations. To expedite universal application of the AeD7L1+2 assay we are committed to openly share the AeD7L1+2 recombinant proteins with other research groups. The AeD7L1+2 proteins used in our assay are stable when kept refrigerated after a four-month testing period, with only a minor decay at room temperature. This supports their simple deployment in resource-scarce settings, especially when developed into a quantitative lateral flow assay which will in turn overcome the need for expensive equipment, such as an ELISA reader spectrophotometer, and inter-laboratory standardization. A limiting factor of our assay is the lack of a seropositivity cutoff, which is challenging to define given that most Cambodians are under constant *Ae. aegypti* biting pressure. Additionally, we did not study other Ab isotypes or IgG subclasses but focused on IgG responses given their abundance and robustness in freeze-and-thaw cycles (43–45). Future mechanistic studies would benefit from in-depth investigations into the humoral and cellular immune response to *Ae. aegypti* saliva to gain understanding of how AeD7L1+2 Abs impact dengue disease outcome. Knowing the rate at which AeD7L Abs decay after disrupting mosquito-human contact will be critical to optimize its use as an epidemiological tool. Abs against saliva of several vector species, including *Ae. aegypti*, are known to decline with age (11, 46–51). Yet, how these dynamics impact dengue disease outcome is not well understood. Discerning these age trends will be crucial when factoring them into the design of future studies. Overall, bridging these gaps will contribute to a more nuanced understanding of the interplay between immunity and *Aedes*-borne diseases and expose potential strategies for their prevention and control.

The highly sensitive and *Aedes*-specific novel AeD7L1+2 exposure assay will be useful in advancing the epidemiological understanding of *Aedes*-borne diseases. Our assay will encourage and facilitate the standardization of future serosurveys and epidemiological studies by its ability to provide a robust estimation of human-mosquito contact in a high-throughput fashion.

## 5 Material and methods

### 5.1 Study population and sample selection

The samples selected for the present study originate from the PAGODAS longitudinal cohort study conducted in Cambodia, of which study protocol and results are reported elsewhere (18, 41). In brief, children enrolled in the study (2–9 years old) were followed up every six months for a period of three years to assess their immune status against dengue and *Ae. aegypti* saliva. Children were considered dengue naïve at study baseline if they were negative on the PanBio Dengue Indirect IgG enzyme-linked immunosorbent assay (ELISA), or, when ELISA positive, they were negative for the presence of DENV-specific neutralizing antibodies (Abs) (n=563). During the follow up period, 41 children developed symptomatic dengue as confirmed by RT-PCR for DENV 1–4. Children who seroconverted via pan-DENV ELISA screening with a corresponding increase in DENV 1-4 neutralization Ab titers over the threshold value of 1:40 at one of the semiannual visits, and who had not reported fever-like symptoms during active febrile surveillance, were considered to have had asymptomatic dengue (n=173).

For the present study, a random sample of baseline-dengue naïve children, stratified by quartiles of anti-SGH Ab response levels (i.e., high, medium, low), were selected for the initial immunogenicity screening of *Ae. aegypti* salivary proteins by Western blotting (n = 102). Ab responses against five identified salivary antigens and combinations thereof were then measured in a random subset of samples collected at all seven visits from 18 baseline-dengue naïve children (n=126) to investigate correlations with the Ab response against whole SGH, and to evaluate the seasonality of these Ab responses. Associations between anti-salivary protein Abs and disease outcome (i.e., no dengue infection, symptomatic disease, or asymptomatic disease) were assessed and correlations confirmed using all baseline-dengue naïve samples (n=563). Negative controls were selected from National Institutes of Health (NIH) Clinical Center (CC) blood donor samples that were non-reactive against *Ae. aegypti* saliva (n = 10), positive controls were selected from human sera samples with high reactivity against *Ae. aegypti* SGH, as defined by ELISA, and stored at the International Centre of Excellence in Research laboratory.

### 5.2 Ethics

The study protocol was approved by the institutional review boards (IRB) at the US National Institutes of Health and the National Ethics Committee on Human Research (NECHR) in Cambodia. The parents or guardians of all pediatric participants provided signed informed consent to participate in the study (ClinicalTrials.gov identifier: NCT03534245). Sera from healthy U.S. blood donors were collected following signed informed consent from participants in the National Institutes of Health Clinical Center IRB-approved protocol entitled *Collection and Distribution of Blood Components from Healthy Donors for In Vitro Research Use* (ClinicalTrials.gov identifier: NCT00001846).

### 5.3 Mosquito rearing, dissection, and salivary gland homogenization

*Ae. aegypti* (Kampong Speu strain, Cambodia and Liverpool strain, Rockville, MD), *Ae. albopictus* (Phnom Penh strain, Cambodia), *Cx. quinquefasciatus* (Phnom Penh strain, Cambodia), and *An. dirus* (Mondulkiri strain, Cambodia) mosquitoes were reared in a controlled environment insectary at the National Center for Parasitology, Entomology and Malaria Control (Cambodia) under a 12-h light-dark cycle, at 26° C, and 75 % relative humidity. *Ae. aegypti* Rockefeller strain was reared under the same conditions at the Laboratory of Malaria and Vector Research (National Institutes of Health, U.S.). Larvae were fed with fish food (Tetra, USA) every 2 days until pupae development, while adults were offered 10 % sucrose solution, *ad libitum*. Salivary glands from females 7 – 10 days post-eclosion were dissected under a stereomicroscope at one gland per microliter of sterile 1 x phosphate-buffered saline (PBS), pH 7.4 (Thermo Scientific) and stored at –80° C. Prior to use, salivary glands were disrupted through ultrasonication, and supernatant collected after removal of insoluble debris by centrifugation at 14,000 rpm for 5 min at 4° C.

### 5.4 Production of recombinant *Ae. aegypti* salivary proteins

The human cell line HEK293 was used to recombinantly express 18 *Ae. aegypti* recombinant proteins, as described previously (Valenzuela-Leon et al., 2022). After identification of the most immunogenic proteins, we scaled up the production of AeApyrase, AeD7L1, AeD7L2, NIH-23, and NIH-27 and codon-optimized them for mammalian expression, synthesized with a carboxyl-terminal HIS-tag (GenScript) and cloned into pHEK293 Ultra Expression Vectors (Takara). The SAIC Advanced Research Facility (Frederick, MD) then transfected the EXPI293F human cells and recombinantly expressed the proteins. Proteins were purified from the cell culture supernatant using AmMag Ni Magnetic beads (GenScript) and the AmMag Wand D12 (GenScript), following manufacturer’s instructions. Proteins dialysis was performed overnight with sterile PBS (Lonza) using 10K MWCO Slide-A-Lyzer Dialysis Cassettes (Thermo Scientific). The BCA Protein Assay Kit (Pierce) was used to quantify the protein concentration, after which SDS-PAGE was performed using NuPAGE Novex 4–12 % Bis-Tris gradient protein gels (Thermo Fisher Scientific) to assess the quality of the purified proteins. Proteins were visualized with Coomassie G-250 (SimplyBlue SafeStain, Invitrogen) using the eStain protein staining device (GenScript), following manufacturer’s instructions.

### 5.5 Enzyme-linked immunosorbent assay

Total anti-*Ae. aegypti* SGH IgG was measured by performing ELISAs as adapted from our previously described protocol (18). In brief, 96-well Immulon plates were coated with either *Ae. aegypti* SGH (2 µg/ml, Kampong Speu strain) or our *Ae. aegypti* recombinant salivary proteins (5 µg/ml) diluted in carbonate-bicarbonate buffer (pH 9.6) and incubated overnight at 4° C. Plates were blocked with 200 μl of TBST/BSA 4 % for 1 hour at RT and incubated with sera (diluted 1:200) for 2 hours at RT. Plates were washed three times with TBST. Secondary anti-human IgG Abs conjugated to alkaline phosphatase were added at a concentration of 1:10,000, diluted in TBST-BSA 4 % (Millipore-Sigma A1543), and incubated for 1 hour at RT. After six washes, Alkaline Phosphatase Yellow (pNPP) substrate was added, and absorbance read at 450 nm within 30 minutes. Sera samples were tested in duplicate and retested if the coefficient of variation was above 20 %. The optical density (OD) value of the blank control, included in each plate, was subtracted from the raw sample OD values. The same set of positive and negative control samples was included in each plate (per antigen) to allow for inter-plate standardization (i.e., OD sample / (OD positive control – OD negative control).

### 5.6 Western blot

Western blot experiments were performed to identify the most immunogenic *Ae. aegypti* salivary proteins. In brief, 50 µg of *Ae. aegypti* SGH (Rockefeller strain) or 5 µg of recombinant proteins were denatured at 70° C for 10 minutes under non-reducing conditions and loaded into one 2D well (NuPAGE™ 4 – 12 % Bis-Tris Gel, Thermo Fisher Scientific) or in each lane of a 10-well gel (Bolt™ 4 – 12 % Bis-Tris Plus, Thermo Fisher Scientific), respectively. Proteins were separated by SDS-PAGE at 200 V for 40 min. The proteins were transferred to a 0.45 µm nitrocellulose membrane (Thermo Fisher Scientific) and blocked with 5 % nonfat dried milk dissolved in Tris-buffered saline with 0.05 % Tween 20 (TBST–5 % milk) (Sigma) at 4° C overnight. The next day, the membrane was incubated with sera samples (diluted 1:200) in TBST–5 % milk. Samples were either individually loaded in the mini-protean II multiscreen apparatus (Bio-Rad) (i.e., for the initial immunogenicity screening of whole SGH) or pooled and transferred to hybridization bags (Kapak SealPAK, ProAmpac) (i.e., to verify the immunogenicity of the five identified proteins). After a 2-hour incubation at RT, the membranes were incubated for 1 hour at RT with secondary alkaline phosphatase-conjugated goat anti-human IgG heavy and light chains (IgG H+L; diluted at 1:10,000) (SIGMA). Between each phase, membranes were washed three times for five minutes with TBST. The chromogenic reaction was initiated with Alkaline Phosphatase Western Blue® Stabilized Substrate (Promega) and stopped with distilled water after approximately 10 to 15 minutes. Images were captured using a KwikQuant Digital Western Blot Detection System (Kindle Biosciences). Kindle Biosciences’ KwikQuant Image Analyzer software was used to analyze images and to identify reactive protein bands. For the Western blot inhibition assay, we first quenched all *Aedes* specific antibodies by incubating the pooled sera with 5 μg of both recombinant AeD7L1 and AeD7L2 in hybridization bags overnight at RT prior to incubation with the membrane.

### 5.7 Phylogenetic analysis

The sequences of the salivary D7 long form proteins from *Ae. aegypti, Ae. albopictus* and *Cx. quinquefasciatus* were aligned after removal of their predicted signal peptide sequence (52). Multiple sequence alignment and identity/similarity matrices were constructed in Geneious v2022.1.1 using the MUSCLE algorithm (53) and the PAM200 scoring matrix. Homologues of AeD7L1 and AeD7L2 were identified in the Non-Redundant (NR) protein database and the Transcriptome Shotgun Assembly (TSA) database using the Basic Local Alignment Search Tool (54). Mosquitoes with the most homologous sequences to *Ae. aegypti* (based on their E-value) were selected. The unrooted evolutionary tree was then constructed with PAUP 4.0 (55) in Geneious v2022.1.1, following the maximum likelihood procedure of the WAG amino acid substitution matrix model. The partial deletion option was applied for gaps/missing data. The reliability of the trees was tested by the bootstrap method (n=1,000).

### 5.8 Statistical analysis

Normality of OD values was assessed by visual inspection of their distribution and formally tested using the Shapiro-Wilk test. A Kruskal-Wallis rank test was used to test for differences among > 2 groups (for non-parametric inferences), with a post-hoc Dunn’s test. A Friedman rank test with a post-hoc Conover-Iman test was used to test for differences among more than two paired groups. Multiple comparisons were adjusted using the Benjamini-Hochberg procedure. The Kendall W was used as a measure of the Friedman test effect size. Changes in immune responses with age and disease outcome were tested by fitting generalized linear models (GLM) to the Ab values, where a gamma distribution and log-link function gave the best fit by log-likelihood goodness-of-fit statistics. An age and disease outcome interaction term was included in the model, with age treated as a mean-centered continuous variable. Correlations between anti-SGH and anti-recombinant protein IgG responses were estimated by the Pearson method to investigate a linear relationship between both responses (r), and by the Spearman’s rank method to assess monotonic relationships (r_s_). Cox proportional hazards (PH) models were used to model time to any dengue infection (in days), comparing those with high vs. low Ab responses at baseline, and controlling for sex, age, insecticide use, larvicide use, bed net use, mosquito coil use, school attendance, socio-economic status, and number of toilets around the house. To fit the Cox PH models, Ab responses were categorized as high or low based on a cutoff established using tertiles of Ab responses: ‘high’ is defined as the middle and highest tertiles, ‘low’ as the lowest tertile. All statistical analyses were performed in R Statistical Software (56), and graphical representations created using the ‘ggplot2’ and the ‘corrplot’ package in R (57, 58).

## Data availability statement

The raw data supporting this article can be found in the supplementary files, Further inquiries can be directed to the corresponding author.

## Ethics statement

The study protocol was approved by the institutional review boards (IRB) at the US National Institutes of Health and the National Ethics Committee on Human Research (NECHR) in Cambodia. The parents or guardians of all pediatric participants provided signed informed consent to participate in the study (ClinicalTrials.gov identifier: NCT03534245). Sera from NIH CC blood donors were collected following signed informed consent from participants in the National Institutes of Health Clinical Center IRB-approved protocol entitled *Collection and Distribution of Blood Components from Healthy Donors for In Vitro Research Use* (protocol number: 99-CC-0168; ClinicalTrials.gov identifier: NCT00001846).

## Author contributions

FO, SC, LW, JGV, JEM, CY, CL, RL, and RH conceptualized the study. SC, LW, SN, PL, KT, JO, RSC, AP, DK, SL, RS performed experiments and discussed data. PCVL and EC discussed data and provided reagents. FO, SC and LW performed data analysis. FO, SC, LW wrote the manuscript. All authors contributed to the article and approved the submitted version.

## Data Availability

All data produced in the present study are available upon reasonable request to the authors

## Acknowledgments

We thank the participating children and their parents in Chbar Mon, Kampong Speu. We appreciate the continuous support and collaboration from the provincial health department and hospital directors of Kampong Speu province. Additionally, we commend the clinical staff of Kampong Speu District Referral Hospital for their outstanding patient care in this project.

## References

1. Bhatt S, Gething PW, Brady OJ, Messina JP, Farlow AW, Moyes CL, Drake JM, Brownstein JS, Hoen AG, Sankoh O, Myers MF, George DB, Jaenisch T, Wint GR, Simmons CP, Scott TW, Farrar JJ, Hay SI. 2013. The global distribution and burden of dengue. Nature 496:504–7.

2. Tian N, Zheng JX, Guo ZY, Li LH, Xia S, Lv S, Zhou XN. 2022. Dengue Incidence Trends and Its Burden in Major Endemic Regions from 1990 to 2019. Trop Med Infect Dis 7.

3. WHO. 2023. Dengue and severe dengue, *on* World Health Organisation. https://www.who.int/news-room/fact-sheets/detail/dengue-and-severe-dengue. Accessed

4. Alied M, Nguyen D, Abdul Aziz JM, Vinh DP, Huy NT. 2023. Dengue fever on the rise in Southeast Asia. Pathog Glob Health 117:1–2.

5. Ribeiro JM. 1987. Role of saliva in blood-feeding by arthropods. Annu Rev Entomol 32:463–78.

6. Dhawan R, Kumar M, Mohanty AK, Dey G, Advani J, Prasad TS, Kumar A. 2017. Mosquito-Borne Diseases and Omics: Salivary Gland Proteome of the Female Aedes aegypti Mosquito. OMICS 21:45–54.

7. Chowdhury A, Modahl CM, Misse D, Kini RM, Pompon J. 2021. High resolution proteomics of Aedes aegypti salivary glands infected with either dengue, Zika or chikungunya viruses identify new virus specific and broad antiviral factors. Sci Rep 11:23696.

8. Schneider CA, Calvo E, Peterson KE. 2021. Arboviruses: How Saliva Impacts the Journey from Vector to Host. Int J Mol Sci 22.

9. Londono-Renteria B, Cardenas JC, Cardenas LD, Christofferson RC, Chisenhall DM, Wesson DM, McCracken MK, Carvajal D, Mores CN. 2013. Use of anti-Aedes aegypti salivary extract antibody concentration to correlate risk of vector exposure and dengue transmission risk in Colombia. PLoS One 8:e81211.

10. Mathieu-Daudé F, Claverie A, Plichart C, Boulanger D, Mphande FA, Bossin HC. 2018. Specific human antibody responses to Aedes aegypti and Aedes polynesiensis saliva: A new epidemiological tool to assess human exposure to disease vectors in the Pacific. PLOS Neglected Tropical Diseases 12.

11. Doucoure S, Mouchet F, Cournil A, Le Goff G, Cornelie S, Roca Y, Giraldez MG, Simon ZB, Loayza R, Misse D, Flores JV, Walter A, Rogier C, Herve JP, Remoue F. 2012. Human antibody response to Aedes aegypti saliva in an urban population in Bolivia: a new biomarker of exposure to Dengue vector bites. Am J Trop Med Hyg 87:504–10.

12. Doucoure S, Mouchet F, Cornelie S, DeHecq JS, Rutee AH, Roca Y, Walter A, Herve JP, Misse D, Favier F, Gasque P, Remoue F. 2012. Evaluation of the human IgG antibody response to Aedes albopictus saliva as a new specific biomarker of exposure to vector bites. PLoS Negl Trop Dis 6:e1487.

13. Doucoure S, Cornelie S, Patramool S, Mouchet F, Demettre E, Seveno M, Dehecq JS, Rutee H, Herve JP, Favier F, Misse D, Gasque P, Remoue F. 2013. First screening of Aedes albopictus immunogenic salivary proteins. Insect Mol Biol 22:411–23.

14. Ndille EE, Dubot-Peres A, Doucoure S, Mouchet F, Cornelie S, Sidavong B, Fournet F, Remoue F. 2014. Human IgG antibody response to Aedes aegypti Nterm-34 kDa salivary peptide as an indicator to identify areas at high risk for dengue transmission: a retrospective study in urban settings of Vientiane city, Lao PDR. Trop Med Int Health 19:576–80.

15. Machain-Williams C, Mammen MP, Jr., Zeidner NS, Beaty BJ, Prenni JE, Nisalak A, Blair CD. 2012. Association of human immune response to Aedes aegypti salivary proteins with dengue disease severity. Parasite Immunol 34:15–22.

16. Etienne V, Gallagher A, Christofferson RC, McCracken MK, Cummings DAT, Long MT. 2023. Antibodies to Aedes spp. salivary proteins: a systematic review and pooled analysis. Frontiers in Tropical Diseases 4.

17. Londono-Renteria BL, Shakeri H, Rozo-Lopez P, Conway MJ, Duggan N, Jaberi-Douraki M, Colpitts TM. 2018. Serosurvey of Human Antibodies Recognizing Aedes aegypti D7 Salivary Proteins in Colombia. Front Public Health 6:111.

18. Manning JE, Chea S, Parker DM, Bohl JA, Lay S, Mateja A, Man S, Nhek S, Ponce A, Sreng S, Kong D, Kimsan S, Meneses C, Fay MP, Suon S, Huy R, Lon C, Leang R, Oliveira F. 2022. Development of Inapparent Dengue Associated With Increased Antibody Levels to Aedes aegypti Salivary Proteins: A Longitudinal Dengue Cohort in Cambodia. J Infect Dis 226:1327–1337.

19. Ribeiro JM, Arca B, Lombardo F, Calvo E, Phan VM, Chandra PK, Wikel SK. 2007. An annotated catalogue of salivary gland transcripts in the adult female mosquito, Aedes aegypti. BMC Genomics 8:6.

20. Valenzuela JG, Pham VM, Garfield MK, Francischetti IM, Ribeiro JM. 2002. Toward a description of the sialome of the adult female mosquito Aedes aegypti. Insect Biochem Mol Biol 32:1101–22.

21. Calvo E, Tokumasu F, Marinotti O, Villeval JL, Ribeiro JMC, Francischetti IMB. 2007. Aegyptin, a novel mosquito salivary gland protein, specifically binds to collagen and prevents its interaction with platelet glycoprotein VI, integrin alpha2beta1, and von Willebrand factor. J Biol Chem 282:26928–26938.

22. Calvo E, Mans BJ, Andersen JF, Ribeiro JM. 2006. Function and evolution of a mosquito salivary protein family. J Biol Chem 281:1935–42.

23. Matthews BJ, Dudchenko O, Kingan SB, Koren S, Antoshechkin I, Crawford JE, Glassford WJ, Herre M, Redmond SN, Rose NH, Weedall GD, Wu Y, Batra SS, Brito-Sierra CA, Buckingham SD, Campbell CL, Chan S, Cox E, Evans BR, Fansiri T, Filipovic I, Fontaine A, Gloria-Soria A, Hall R, Joardar VS, Jones AK, Kay RGG, Kodali VK, Lee J, Lycett GJ, Mitchell SN, Muehling J, Murphy MR, Omer AD, Partridge FA, Peluso P, Aiden AP, Ramasamy V, Rasic G, Roy S, Saavedra-Rodriguez K, Sharan S, Sharma A, Smith ML, Turner J, Weakley AM, Zhao Z, Akbari OS, Black WCt, Cao H, et al. 2018. Improved reference genome of Aedes aegypti informs arbovirus vector control. Nature 563:501–507.

24. Martins M, Ramos LFC, Murillo JR, Torres A, de Carvalho SS, Domont GB, de Oliveira DMP, Mesquita RD, Nogueira FCS, Maciel-de-Freitas R, Junqueira M. 2021. Comprehensive Quantitative Proteome Analysis of Aedes aegypti Identifies Proteins and Pathways Involved in Wolbachia pipientis and Zika Virus Interference Phenomenon. Front Physiol 12:642237.

25. Wasinpiyamongkol L, Patramool S, Luplertlop N, Surasombatpattana P, Doucoure S, Mouchet F, Seveno M, Remoue F, Demettre E, Brizard JP, Jouin P, Biron DG, Thomas F, Misse D. 2010. Blood-feeding and immunogenic Aedes aegypti saliva proteins. Proteomics 10:1906–16.

26. Champagne DE, Smartt CT, Ribeiro JM, James AA. 1995. The salivary gland-specific apyrase of the mosquito Aedes aegypti is a member of the 5’-nucleotidase family. Proc Natl Acad Sci U S A 92:694–8.

27. Sagna AB, Kassie D, Couvray A, Adja AM, Hermann E, Riveau G, Salem G, Fournet F, Remoue F. 2019. Spatial Assessment of Contact Between Humans and Anopheles and Aedes Mosquitoes in a Medium-Sized African Urban Setting, Using Salivary Antibody-Based Biomarkers. J Infect Dis 220:1199–1208.

28. Choi Y, Tang CS, McIver L, Hashizume M, Chan V, Abeyasinghe RR, Iddings S, Huy R. 2016. Effects of weather factors on dengue fever incidence and implications for interventions in Cambodia. BMC Public Health 16:241.

29. Wai KT, Arunachalam N, Tana S, Espino F, Kittayapong P, Abeyewickreme W, Hapangama D, Tyagi BK, Htun PT, Koyadun S, Kroeger A, Sommerfeld J, Petzold M. 2012. Estimating dengue vector abundance in the wet and dry season: implications for targeted vector control in urban and peri-urban Asia. Pathog Glob Health 106:436–45.

30. Parker DM, Medina C, Bohl J, Lon C, Chea S, Lay S, Kong D, Nhek S, Man S, Doehl JSP, Leang R, Kry H, Rekol H, Oliveira F, Minin VM, Manning JE. 2023. Determinants of exposure to Aedes mosquitoes: A comprehensive geospatial analysis in peri-urban Cambodia. Acta Trop 239:106829.

31. Fustec B, Phanitchat T, Aromseree S, Pientong C, Thaewnongiew K, Ekalaksananan T, Cerqueira D, Poinsignon A, Elguero E, Bangs MJ, Alexander N, Overgaard HJ, Corbel V. 2021. Serological biomarker for assessing human exposure to Aedes mosquito bites during a randomized vector control intervention trial in northeastern Thailand. PLoS Negl Trop Dis 15:e0009440.

32. Elanga Ndille E, Doucoure S, Poinsignon A, Mouchet F, Cornelie S, D’Ortenzio E, DeHecq JS, Remoue F. 2016. Human IgG Antibody Response to Aedes Nterm-34kDa Salivary Peptide, an Epidemiological Tool to Assess Vector Control in Chikungunya and Dengue Transmission Area. PLoS Negl Trop Dis 10:e0005109.

33. Doucoure S, Mouchet F, Cornelie S, Drame PM, D’Ortenzio E, DeHecq JS, Remoue F. 2014. Human antibody response to Aedes albopictus salivary proteins: a potential biomarker to evaluate the efficacy of vector control in an area of Chikungunya and Dengue Virus transmission. Biomed Res Int 2014:746509.

34. Valenzuela JG, Charlab R, Gonzalez EC, de Miranda-Santos IK, Marinotti O, Francischetti IM, Ribeiro JM. 2002. The D7 family of salivary proteins in blood sucking diptera. Insect Mol Biol 11:149–55.

35. Alvarenga PH, Andersen JF. 2022. An Overview of D7 Protein Structure and Physiological Roles in Blood-Feeding Nematocera. Biology (Basel) 12.

36. Martin-Martin I, Kojin BB, Aryan A, Williams AE, Molina-Cruz A, Valenzuela-Leon PC, Shrivastava G, Botello K, Minai M, Adelman ZN, Calvo E. 2023. Aedes aegypti D7 long salivary proteins modulate blood feeding and parasite infection. mBio doi:10.1128/mbio.02289-23:e0228923.

37. Oktarianti R, Senjarini K, Hayano T, Fatchiyah F, Aulanni’am. 2015. Proteomic analysis of immunogenic proteins from salivary glands of Aedes aegypti. J Infect Public Health 8:575–82.

38. Oseno B, Marura F, Ogwang R, Muturi M, Njunge J, Nkumama I, Mwakesi R, Mwai K, Rono MK, Mwakubambanya R, Osier F, Tuju J. 2022. Characterization of Anopheles gambiae D7 salivary proteins as markers of human-mosquito bite contact. Parasit Vectors 15:11.

39. Leta S, Beyene TJ, De Clercq EM, Amenu K, Kraemer MUG, Revie CW. 2018. Global risk mapping for major diseases transmitted by Aedes aegypti and Aedes albopictus. Int J Infect Dis 67:25–35.

40. Kraemer MU, Sinka ME, Duda KA, Mylne AQ, Shearer FM, Barker CM, Moore CG, Carvalho RG, Coelho GE, Van Bortel W, Hendrickx G, Schaffner F, Elyazar IR, Teng HJ, Brady OJ, Messina JP, Pigott DM, Scott TW, Smith DL, Wint GR, Golding N, Hay SI. 2015. The global distribution of the arbovirus vectors Aedes aegypti and Ae. albopictus. Elife 4:e08347.

41. Manning JE, Oliveira F, Parker DM, Amaratunga C, Kong D, Man S, Sreng S, Lay S, Nang K, Kimsan S, Sokha L, Kamhawi S, Fay MP, Suon S, Ruhl P, Ackerman H, Huy R, Wellems TE, Valenzuela JG, Leang R. 2018. The PAGODAS protocol: pediatric assessment group of dengue and Aedes saliva protocol to investigate vector-borne determinants of Aedes-transmitted arboviral infections in Cambodia. Parasit Vectors 11:664.

42. Conway MJ, Londono-Renteria B, Troupin A, Watson AM, Klimstra WB, Fikrig E, Colpitts TM. 2016. Aedes aegypti D7 Saliva Protein Inhibits Dengue Virus Infection. PLoS Negl Trop Dis 10:e0004941.

43. Rastawicki W, Smietanska K, Rokosz N, Jagielski M. 2012. [Effect of multiple freeze-thaw cycles on detection of IgA, IgG and IgM antibodies to selected bacterial antigens]. Med Dosw Mikrobiol 64:79–85.

44. Pinsky NA, Huddleston JM, Jacobson RM, Wollan PC, Poland GA. 2003. Effect of multiple freeze-thaw cycles on detection of measles, mumps, and rubella virus antibodies. Clin Diagn Lab Immunol 10:19–21.

45. Shurrab FM, Al-Sadeq DW, Amanullah F, Younes SN, Al-Jighefee H, Younes N, Dargham SR, Yassine HM, Abu Raddad LJ, Nasrallah GK. 2021. Effect of multiple freeze-thaw cycles on the detection of anti-SARS-CoV-2 IgG antibodies. J Med Microbiol 70.

46. Peng Z, Ho MK, Li C, Simons FE. 2004. Evidence for natural desensitization to mosquito salivary allergens: mosquito saliva specific IgE and IgG levels in children. Ann Allergy Asthma Immunol 93:553–6.

47. Oliveira F, Traore B, Gomes R, Faye O, Gilmore DC, Keita S, Traore P, Teixeira C, Coulibaly CA, Samake S, Meneses C, Sissoko I, Fairhurst RM, Fay MP, Anderson JM, Doumbia S, Kamhawi S, Valenzuela JG. 2013. Delayed-type hypersensitivity to sand fly saliva in humans from a leishmaniasis-endemic area of Mali is Th1-mediated and persists to midlife. J Invest Dermatol 133:452–9.

48. Buezo Montero S, Gabrieli P, Montarsi F, Borean A, Capelli S, De Silvestro G, Forneris F, Pombi M, Breda A, Capelli G, Arca B. 2020. IgG Antibody Responses to the Aedes albopictus 34k2 Salivary Protein as Novel Candidate Marker of Human Exposure to the Tiger Mosquito. Front Cell Infect Microbiol 10:377.

49. Cardenas JC, Drame PM, Luque-Burgos KA, Berrio JD, Entrena-Mutis E, Gonzalez MU, Carvajal DJ, Gutierrez-Silva LY, Cardenas LD, Colpitts TM, Mores CN, Londono-Renteria B. 2019. IgG1 and IgG4 antibodies against Aedes aegypti salivary proteins and risk for dengue infections. PLoS One 14:e0208455.

50. Montiel J, Carbal LF, Tobon-Castano A, Vasquez GM, Fisher ML, Londono-Renteria B. 2020. IgG antibody response against Anopheles salivary gland proteins in asymptomatic Plasmodium infections in Narino, Colombia. Malar J 19:42.

51. Willen L, Milton P, Hamley JID, Walker M, Osei-Atweneboana MY, Volf P, Basanez MG, Courtenay O. 2022. Demographic patterns of human antibody levels to Simulium damnosum s.l. saliva in onchocerciasis-endemic areas: An indicator of exposure to vector bites. PLoS Negl Trop Dis 16:e0010108.

52. Almagro Armenteros JJ, Tsirigos KD, Sonderby CK, Petersen TN, Winther O, Brunak S, von Heijne G, Nielsen H. 2019. SignalP 5.0 improves signal peptide predictions using deep neural networks. Nat Biotechnol 37:420–423.

53. Edgar RC. 2004. MUSCLE: a multiple sequence alignment method with reduced time and space complexity. BMC Bioinformatics 5:113.

54. Altschul SF, Gish W, Miller W, Myers EW, Lipman DJ. 1990. Basic local alignment search tool. J Mol Biol 215:403–10.

55. Wilgenbusch JC, Swofford D. 2003. Inferring evolutionary trees with PAUP*. Curr Protoc Bioinformatics Chapter 6:Unit 6 4.

56. R Core Team. 2023. R: A language and environment for statistical computing., vv4.3.1. R Foundation for Statistical Computing, http://www.R-project.org.

57. Wickham H. 2016. ggplot2: Elegant Graphics for Data Analysis. Springer-Verlag New York.

58. Wei T, Simko V. 2021. R package ‘corrplot’: Visualization of a Correlation Matrix. https://github.com/taiyun/corrplot. Accessed

