## SupplementaryMaterial for "Antibodies to *Aedes aegypti* D7L salivary proteins as a new serological tool to estimate human exposure to *Aedes* mosquitoes"

### 1. Supplementary Figures and Tables

#### a. Supplementary Figures

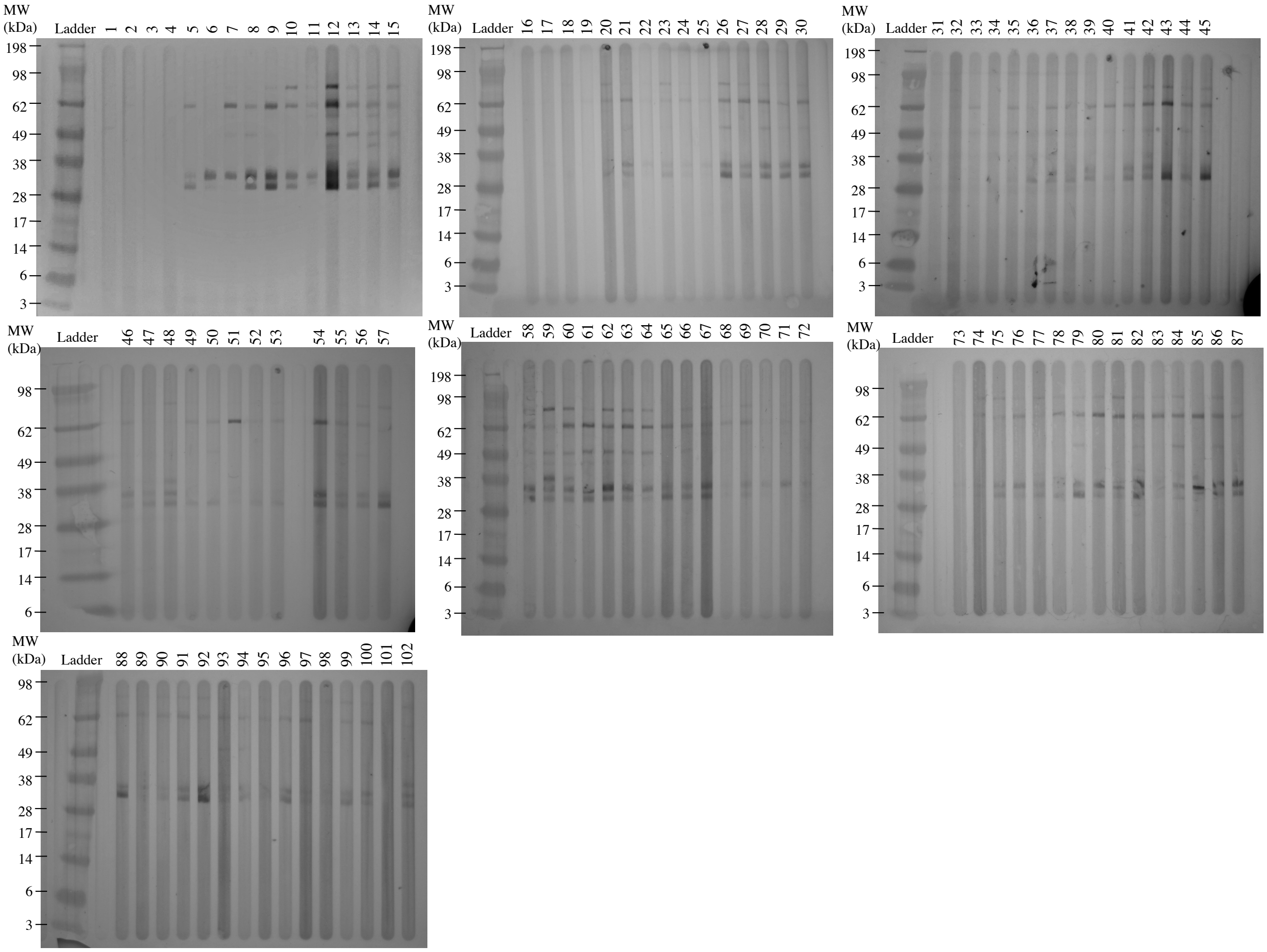


**Supplementary Figure 1. Western blots for reactivity of Cambodian sera with whole salivary gland homogenate of *Ae. aegypti*.** Each lane represents the serum reactivity of one Cambodian individual with SGH. MW: Molecular Weight; SGH: Salivary Gland Homogenate.

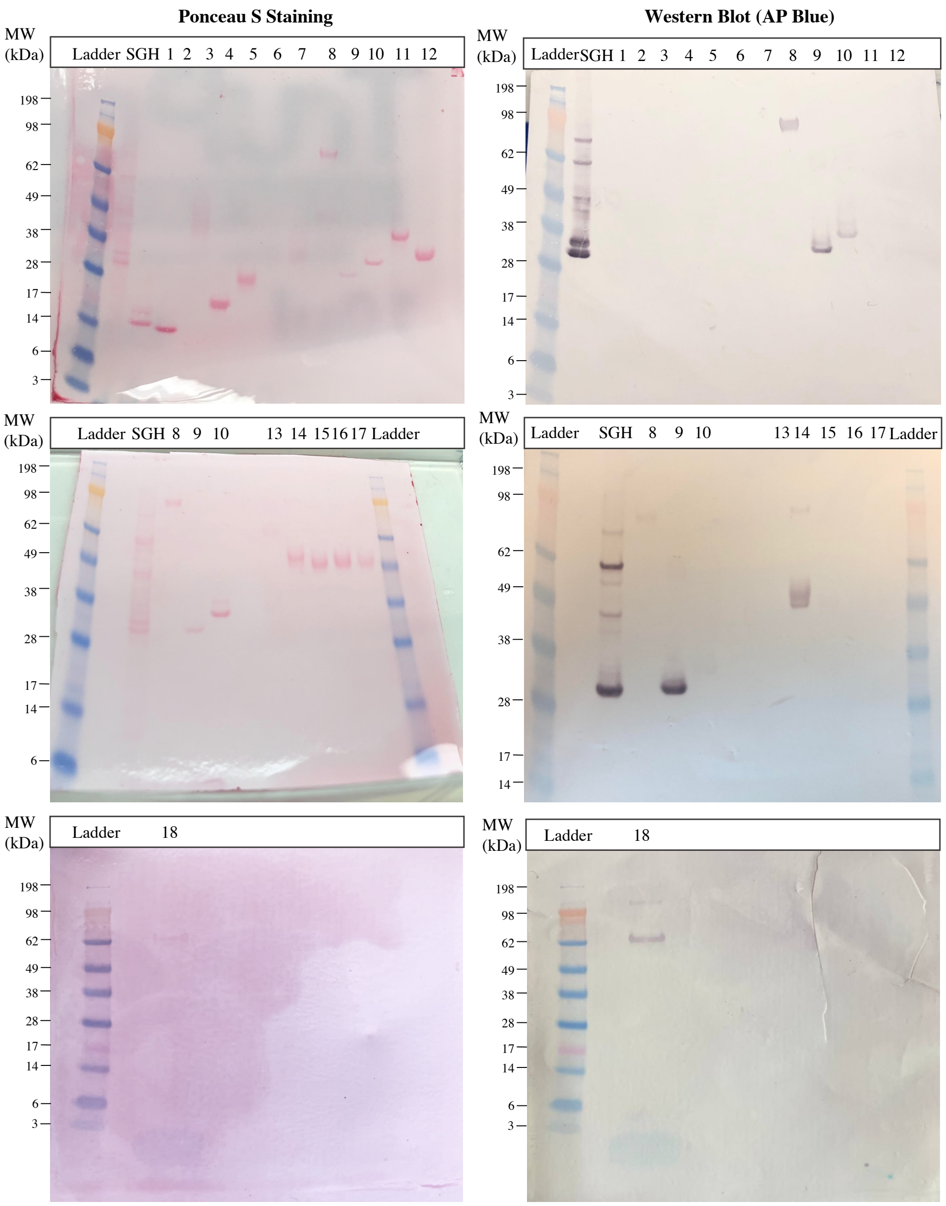


| Lane number | Protein Name | Estimated MW (kDA) |
|---|---|---|
| 1 | NIH435-36 | 20.05 |
| 2 | NIH435-8 | 20.6 |
| 3 | NIH435-4 | 21 |
| 4 | NIH435-29 | 21.16 |
| 5 | NIH435-35 | 21.25 |
| 6 | Aegyptin | 30 |
| 7 | NIH435-32 | 36.4 |
| 8 | NIH435-27 | 36.7 |
| 9 | AeD7L1 | 37 |
| 10 | AeD7L2 | 37 |
| 11 | NIH435-31 | 38.721 |
| 12 | NIH435-24 | 39.13 |
| 13 | NIH435-16 | 20.05 |
| 14 | NIH435-23 | 20.6 |
| 15 | NIH435-19 | 21 |
| 16 | NIH435-6 | 21.16 |
| 17 | NIH435-21 | 21.25 |
| 18 | AeApyrase | 61.36 |

**Supplementary Figure 2. Ponceau staining and Western Blot of 17 recombinantly expressed *Ae. aegypti* salivary proteins.** Protein names and their estimated molecular weight are shown in the table. MW: Molecular Weight; SGH: Salivary Gland Homogenate.

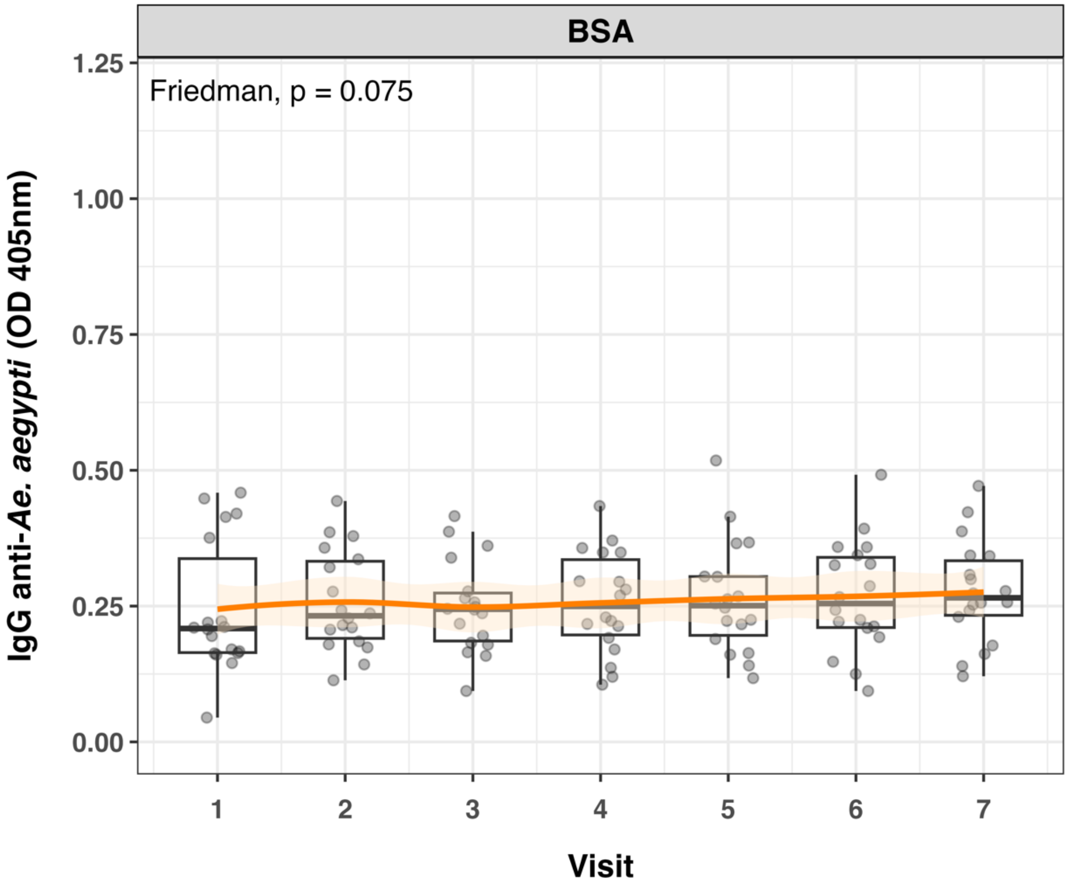


**Supplementary Figure 3.** No significant differences were observed for the Ab responses against BSA during the wet and dry seasons.

#### 2. Supplementary Tables

**Supplementary Table 1. Correlation coefficients, *p* values and confidence intervals for Pearson and Spearman correlation between SGH and all tested combinations of *Ae. aegypti* recombinant salivary proteins.** BSA: Bovine Serum Albumin; CI: Confidence Interval.

| | Pearson Correlation | | | Spearman Correlation | | |
|---|---|---|---|---|---|---|
| Recombinant protein | Coefficient (r) | *p* value | 0.95 CI | Coefficient ($r_s$) | *p* value | 0.95 CI |
| AeD7L1+2 | 0.91 | 4.26e-049 | 0.87 – 0.94 | 0.92 | 4.96e-52 | 0.89 – 0.94 |
| AeD7L1 | 0.82 | 8.75e-32 | 0.75 – 0.87 | 0.84 | 1.00e-34 | 0.78 – 0.89 |
| AeD7L2 | 0.76 | 5.93e-25 | 0.67 – 0.83 | 0.82 | 1.70e-31 | 0.75 – 0.87 |
| AeApyrase + AeD7L1+2 + NIH-27 + NIH-23 | 0.81 | 8.24e-31 | 0.74 – 0.86 | 0.87 | 7.95e-41 | 0.83 – 0.91 |
| AeApyrase + AeD7L1+2 + NIH-27 | 0.84 | 2.12e-34 | 0.78 – 0.88 | 0.90 | 8.98e-48 | 0.87 – 0.93 |
| AeApyrase + AeD7L1+2 + NIH-23 | 0.79 | 1.46e-27 | 0.71 – 0.84 | 0.85 | 4.05e-37 | 0.80 – 0.90 |
| AeApyrase + NIH-23 + NIH-27 | 0.72 | 1.68e-21 | 0.625 – 0.7957 | 0.78 | 2.03e-27 | 0.71 – 0.84 |
| AeD7L1+2 + NIH-27 + NIH-23 | 0.90 | 1.20e-45 | 0.86 – 0.93 | 0.93 | 1.03e-56 | 0.91– 0.95 |
| AeD7L1+2 + NIH-27 | 0.90 | 3.73e-46 | 0.86 – 0.93 | 0.93 | 1.14e-56 | 0.91– 0.95 |
| BSA | 0.17 | 0.051 | -0.001 – 0.34 | 0.15 | 0.10 | -0.03 – 0.31 |

**Supplementary Table 2. Hazard ratios, confidence intervals and *p* values per risk factor tested for dengue seroconversion using Cox regression.** *Aedes aegypti* exposure was estimated using anti-SGH antibody levels. SGH: Salivary Gland Homogenate; HR: Hazard Ratio; CI: Confidence interval.

| Risk factor | HR [CI] | *p* |
|---|---|---|
| ***Aedes aegypti*** **SGH salivary protein antibodies** (*ref: low*) | | |
| *High* | 1.3 [0.94-1.79] | 0.112 |
| **Sex** *(ref: female)* | | |
| *Male* | 1.04 [0.78-1.41] | 0.777 |
| **Age** (per year) | 1.08 [0.98-1.19] | 0.132 |
| **Educational status** *(ref: in school)* | | |
| *Not in school* | 0.83 [0.53-1.29] | 0.402 |
| **Socioeconomic class** *(ref: Lower/ very poor)* | | |
| *Middle/upper* | 0.99 [0.67-1.45] | 0.950 |
| **No. of domestic water containers at home** | 0.10 [0.94-1.06] | 0.927 |
| **No. of toilets in the home** | 1.18 [0.97-1.42] | 0.094 |
| **Use of bednets** *(ref: rarely/never)* | | |
| *Regularly/ all of the time* | 0.89 [0.57-1.39] | 0.610 |
| **Use of insecticide spray** *(ref: does not use)* | | |
| *Uses insecticide spray* | 0.69 [0.50-0.94] | 0.020 |
| **Use of larvicide** *(ref: applies larvicide to water)* | | |
| *Does not apply larvicide to water* | 1.22 [0.81-1.83] | 0.349 |
| **How often mosquito coils burned** *(ref: daily/often)* | | 0.023 |
| *Never* | 1.28 [0.91-1.79] | 0.154 |
| *Sometimes/rarely (1-3 times/wk)* | 0.75 [0.49-1.16] | 0.195 |

**Supplementary Table 3. Hazard ratios, confidence intervals and *p* values per risk factor tested for dengue seroconversion using Cox regression.** *Aedes aegypti* exposure was estimated using anti-D7L1+2 antibody levels. HR: Hazard Ratio; CI: Confidence interval.

| Risk factor | HR [CI] | *p* |
|---|---|---|
| ***Aedes aegypti*** **D7L1+2 salivary protein antibodies** (*ref: low*) | | |
| *High* | 1.2 [1.4-0.9] | 0.341 |
| **Sex** *(ref: female)* | | |
| *Male* | 1 [2.4-0.8] | 0.872 |
| **Age** (per year) | 1.1 [1.2-1] | 0.157 |
| **Educational status** *(ref: in school)* | | |
| *Not in school* | 0.8 [1.5-0.5] | 0.391 |
| **Socioeconomic class** *(ref:* L*ower/ very poor)* | | |
| *Middle/upper* | 1 [2.5-0.7] | 0.914 |
| **No. of domestic water containers at home** | 1 [2.6-0.9] | 0.972 |
| **No. of toilets in the home** | 1.2 [1.1-1] | 0.095 |
| **Use of bednets** *(ref: rarely/never)* | | |
| *Regularly/ all of the time* | 0.9 [1.9-0.6] | 0.617 |
| **Use of insecticide spray** *(ref: does not use)* | | |
| *Uses insecticide spray* | 0.7 [1-0.5] | 0.020 |
| **Use of larvicide** *(ref: applies larvicide to water)* | | |
| *Does not apply larvicide to water* | 1.2 [1.4-0.8] | 0.285 |
| **How often mosquito coils burned** *(ref: daily/often)* | | 0.023 |
| *Never* | 1.3 [1.2-0.9] | 1.365 |
| *Sometimes/rarely (1-3 times/wk)* | 0.7 [1.2-0.5] | <0.0001 |
